# Validation of the patient reported outcome measures tool “Catquest” in Odia language

**DOI:** 10.1101/2024.11.02.24316550

**Authors:** Saswati Sen, Matuli Das, AK Kavitha, Aiswariya Mohanty, Anjali Goel

## Abstract

**Aim:** To validate Catquest questionnaire in Odia speaking population of Eastern India

**Methodology:** Prospective study was conducted on 40 patients planned for cataract surgery in a tertiary care centre in eastern India. Demographic data was collected and comprehensive ocular examination which included slit lamp examination, fundus examination, intraocular pressure measurement was done. Patients were asked to fill the Catquest Questionnaire before and six weeks after the surgery. The English version of the questionnaire was translated and validated into Odia for use in the study. Patients filled both Odia and English questionnaires on both occasions. Statistical analysis was done to note the agreement between the answers given in both languages to validate the questionnaire in Odia

**Results:** The ICC values for various activities such as Reading Papers, Seeing to Walk on Uneven Ground, Watching Television, and Preferred Hobby showed perfect reliability, with ICC = 1 both pre-OP and post-OP. This indicates consistent perceptions of these activities across the two time points. Activities such as seeing prices, seeing to do needle work, reading news papers and shopping also showed good reliability. For Reading Text on Television, the ICC values were -0.0588 pre-OP and 0 post-OP. The negative ICC pre-OP indicates very poor reliability or inconsistency, while the zero ICC post-OP suggests a lack of variability.

**Conclusion:** with growing emphasis on patient reported outcomes, both clinical and subjective parameters need to be considered for the success of any procedure. The study shows good correlation between original and translated version and can be incorporated in clinical practice successfully

## Introduction

Cataract is the most common cause of avoidable blindness worldwide. [1] The surgical treatment for cataract has highly improved to offer better results both objectively and subjectively. Post-surgery, the objective improvement can be documented by assessing of visual acuity(VA), intraocular pressure(IOP) or residual refractive error apart from the clinical assessment of anterior and posterior segment of the eye. Subjective assessment or satisfaction post surgery on the other hand includes tools such as robust questionnaires that may effectively report patient’s perception before and after the surgery.[2] Several such tools have been devised and modified with time to assess the patient perception.[3,4] Catquest questionnaire is one such questionnaire designed to assess the patient reported outcome measures related to cataract surgeries.[5] The success of surgery in the current scenario not only depends on the post-operative visual acuity but also on the ability of the person to carry out certain activities post-surgery. It also depends on whether the patient is able to overcome certain perceived problems or disabilities that they faced prior to the surgery. Catquest questionnaire has a set of questions which try to assess the subjective improvement by asking questions on perceived disabilities which may affect one or more vision related function. It is a closed ended questionnaire which helps in better evaluation of the level of subjective improvement in our patients. Validating the questionnaire helps patient’s who speak local language understand the questions better. This in turn will give a better perception of the patient satisfaction as well as the help the surgeon understand what problems patient’s face on a day to day basis. [6] The present study aims to use this questionnaire in the native language and find whether it is relevant in assessing the patient reported outcome in the developing world. This precludes the need to frame new questions and help in better comparison between studies using the same questionnaire.

## Materials and Methods

This was a prospective study conducted in a tertiary care hospital after obtaining Institutional Ethical Committee clearance (KIIT/KIMS/IEC/952). The study complied to the principles of Declaration of Helsinki. The study period was from June 2023 to December 2023.Purposive sampling was used for the study. A total of 40 patients were included in the study. These were patients reporting to ophthalmology outpatient department with operable cataract who underwent evaluation and surgery for cataract in the department of Ophthalmology of a tertiary care centre in Eastern Odisha. Eligible patients were identified to be included in the study and written informed consent was taken. Demographic profile of all the patients was recorded and a thorough clinical history and examination on slit lamp was done. All patients included in the study were bilingual.

Patients with age above 40 years and having cataract with NS grade 2 and above as per LOCS III grading were included in the study. Patients who refused consent for participation in study, having any eye disease causing profound vision loss other than brown cataract such as optic nerve damage, retinal diseases, any patient having pre-existing systemic or ocular disease causing visual discomfort or visual loss other than cataract were excluded. All eyes of the patients in the study underwent a thorough ophthalmic examination as pre-operative requisites for cataract surgery –i.e Visual acuity testing by Snellen’s chart and logMAR chart, Slit lamp biomicroscopy for anterior segment examination, colour vision and contrast sensitivity was also recorded for each patient, dilated fundoscopic examination with +90 D lens and slit lamp bio- microscopy was done and also with +20 D lens and indirect ophthalmoscopy of posterior segment to rule out any other causes of visual defects such as retinopathies or retinal detachment, Intraocular pressure(IOP) was measured using Goldman Applanation Tonometry . Catquest Questionnaires in both English and Odia was administered to the patients, both before and 4 weeks after the surgery. Data from all the completed forms were collected and analyzed.

### Process of Translation and validation

The validated English version of the Catquest questionnaire available in the public domain was first translated into English by two independent translators. (Appendix 1) Both the translators were bilingual and could both read and write in English and Odia. After careful discussion and consideration, a consensus was reached on a single agreed version to be used in the study. The panel decided to use simple Odia terms which would be understandable by the layman in order to understand the nature of the questions. The Odia version was also reviewed by few monolingual members and their suggestions were taken into account before further modification of the questionnaire. This Odia version was then again backtranslated into English. The English version of the validated Catquest questionnaire and the backtranslated English version were again compared by our experts. A few Odia words and phrases were chosen to be changed or modified. Some other words like ‘suikama’, hobby, embroidery, headlight were used as such but not much discrepancy was observed in understanding the essence of the questions by the patients. After confirming that the meaning and language has not been altered, a final Odia version was developed and approved by the expert committee. This Odia version was then used in the study.

### Statistical Analysis

The Intra-Class Correlation Coefficient (ICC) values were calculated using a one-way random effects model with the agreement type to assess the consistency of measurements across different time points. ICC measures the degree of consistency or reliability of measurements across different time points or conditions, with values ranging from 0 to 1, and potentially negative values indicating very poor reliability. The tables in the study were then framed where Intra- Class Correlation Coefficient (ICC) values for various activities and perceived problems both before (pre-OP) and after (post-OP) the operation were derived.

## Results

40 patients were included in the study which was prospective in nature. The majority of patients were of age more than 60 years. There was no significant difference in gender with equal number of males and females being operated for cataract. On comparison, more patients had an educational qualification of post-graduate level or higher. In addition more number of patients were above poverty line (25) as compared to those below poverty line (15). But with respect to gender there was no statistical difference between these demographic parameters. This meant that equal proportion pf patients of both genders had similar educational and financial status. But when it came to surgical procedure, there was significant difference in the surgical procedure conducted between both the genders. Maximum male patients underwent phacoemulsification while females underwent small incision cataract surgery. The details are listed in Table 1.

**Table 1.** Descriptive summary of study participants

| Variables | Male n(%) | Female n(%) | Total (N=40) | p-value |
| --- | --- | --- | --- | --- |
| Age categories |  |  |  |  |
| <60years | 2(10%) | 5 (25%) | 7(17.5%) | 0.211 |
| >60years | 18(90%) | 15 (75%) | 33(82.5%) |  |
| Education |  |  |  |  |
| Graduate (12<sup>th</sup> std pass) | 9(45%) | 6(30%) | 15(37.5%) | 0.327 |
| Post graduate | 11(55%) | 14(70%) | 25(62.5%) |  |
| Income |  |  |  |  |
| Above poverty line | 11(55%) | 14(70%) | 25(62.5%) | 0.327 |
| Below poverty line | 9(45%) | 6(30%) | 15(37.5%) |  |
| Surgical procedure |  |  |  |  |
| Phaco | 19(95%) | 5(25%) | 20(50%) | < 0.00001 |
| SICS | 1(5%) | 15(75%) | 16(40%) |  |
| Total | 20(50%) | 20(50%) |  |  |

The questionnaire had three subsections and 18 items in total with scope for multidimensional answers. Here, lower figures i.e one represented degree of lower disability where as higher figures represented a degree of higher disability. The questions represented disability based on cataract symptoms, day to day activities and preferred hobbies which are affected by the disease. Though the answers to each question were evaluated separately, seven items were broadly categorized and evaluated. (Table 2) The questionnaire was administered to the patients both before and then four weeks after the surgical procedure. For items like reading papers, recognizing faces and seeing to walk on uneven ground Intra class co-efficient was one pre- operatively which showed good understanding of the patients for questions pertaining to these activities in both languages. Post-operatively, except for recognizing faces, other two categories showed good intra class co-efficient. The patients due to a significant improvement of vision were able to recognize faces without any problems. Perhaps that was why the question did not seem very relevant post-operatively in a broad sense. May be questions about finer visual acuity assessment would have judged the parameter better in our setting. Other activities like reading prices on shopping items, doing needlework or engaging in a preferred hobby showed good intra class co-efficient both pre-operatively and post-operatively in both languages indicating that the questions had similar interpretation in both languages. The ICC for Doing Needlework was 0.271 both pre-OP and post-OP, indicating low reliability or consistency in the perception of problems related to this activity, with no improvement observed after the operation.

**Table 2.** Co-relation between Odia and English Questionnaire comparing day to day activities

| Items | Intra Class Corelation Coefficient(ICC) |  |
| --- | --- | --- |
|  | Pre-op | Post-op |
| Reading Papers | 1.00 | 1.00 |
| Recognizing Faces | 1.00 | NA |
| Seeing<br>Prices/Shopping | 0.814 | 1.00 |
| Seeing to walk on<br>uneven ground | 1.00 | 1.00 |
| Seeing to do<br>needlework | 0.88 | 1.00 |
| Reading text on<br>television | 0.21 | 0.21 |
| Seeing to perform a<br>preferred activity | 0.87 | 1 |

The only category which showed lower intra class coefficient values was reading text on television. On analyzing it was found that some terms used in the Odia questionnaire were confusing for the patients. The term “Dooradarshana” and “TV” were used interchangeably. Post these changes, the responses improved. Another reason for this was, many patients were not prescribed correction post surgery which may have led to them skipping the question altogether or giving erronneous responses. The zero ICC post-OP is attributed to the fact that all patients reported no difficulty with reading text on television after the operation, resulting in no variability to measure agreement against.

Overall, the results indicated good agreement between the questionnaires in both languages and was understandable to the local population.

## Discussion

Patient satisfaction is paramount after every surgery. In Ophthalmology even after a simple cataract surgery resulting in excellent vision and other objective parameters, the patients may still not be satisfied.[7] With changing treatment perspective, the use of questionnaires has become desirable to assess the subjective parameters and Catquest questionnaire has been used extensively for the same in various countries.[8] Apart from Catquest several other questionnaires have also been used similarily.[9,10]Though the questions per se may be different but the overall aim and structure is to evaluate visual disability and assess the patient reported outcomes pre and post-surgery.

The aim of the present study was to validate the Catquest questionnaire in the native language so that the patient perspective is easily understandable in their own terms. This included trying to use as much as possible local terms used in day to day practice so that understanding of the questions as well as expression of answers is better for the subjects. Translation improves communication with patients and increases the reach amongst the study population.

Communication in local language also builds trust between the health care professional and the patients. [11] Further validation of the paper ensures that the content is correct as far as the context is concerned. This means that patient problems can be correctly identified and is not ‘lost in translation”. [12,13]

The questions used in Catquest questionnaire are open ended questions divided into three sections which involved questions on daily activities, difficulty in performing those activities and questions on general health. The questionnaire itself and its modified form have been translated and validated in several languages. [14,15,16,17] These questions could be extrapolated to our socio- demographic scenario as well which will give us an insight into the patient’s perspective as well. Hence, the first step was to translate the questions into native language. This process included the use of colloquial terms rather than literary words which made understanding of the paper easier. Though initially the volunteers did help the patient’s in understanding the process of answering the questions but at the end due care was taken so that it was the patient’s own interpretation at the end which was to be recorded. [18,19]

The subjects in the present study did not have any significant socioeconomic disparity except for the choice of surgery in case of males and females. This may be explained by the fact that phacoemulsification in brown cataracts warrants more caution as in comparison to SICS and males may be more pre-disposed to take risks instead of females in our society. [20] But the Catquest questionnaire comprised of questions related to day to day activity which were basically gender –neutral. Hence, they were not biased by the nature of lifestyle the patient’s led.

All the activities like needle work, reading prices on items, reading newspapers were well understood and interpreted in both languages. But question pertaining to watching television showed no variability post op. This was probably due to some problem in interpreting the words used which were then modified and later confirmed to be understandable by the study subjects.

Responses to questions may also be guided by the fact that people who perform some activities more may be less likely to say they have difficulties in performing the task owing to greater adaptability. [21] A previous study using Catquest in Indian population had found the need to alter some questions to cater to the Indian scenario. [22] But the present study showed good co-relation and agreement for the questions in Odia language. English is not yet used as first language in many parts of the world. Translation and validation of these questionnaires may improve doctor-patient communication. This ensures improved empathy, helps in keeping up with the current trends and better patient care as well. [23,24]

## Conclusion

The present study has successfully translated the Catquest Questionnaire in Odia language. This will ensure combining both clinical objectives and subjective parameters to address the patient expectations and concerns. [25] It can be incorporated in clinical practice to ensure better patient care.

## Data Availability

All data produced are contained in the manuscript

## Appendix 1-Catquest Questionnaire

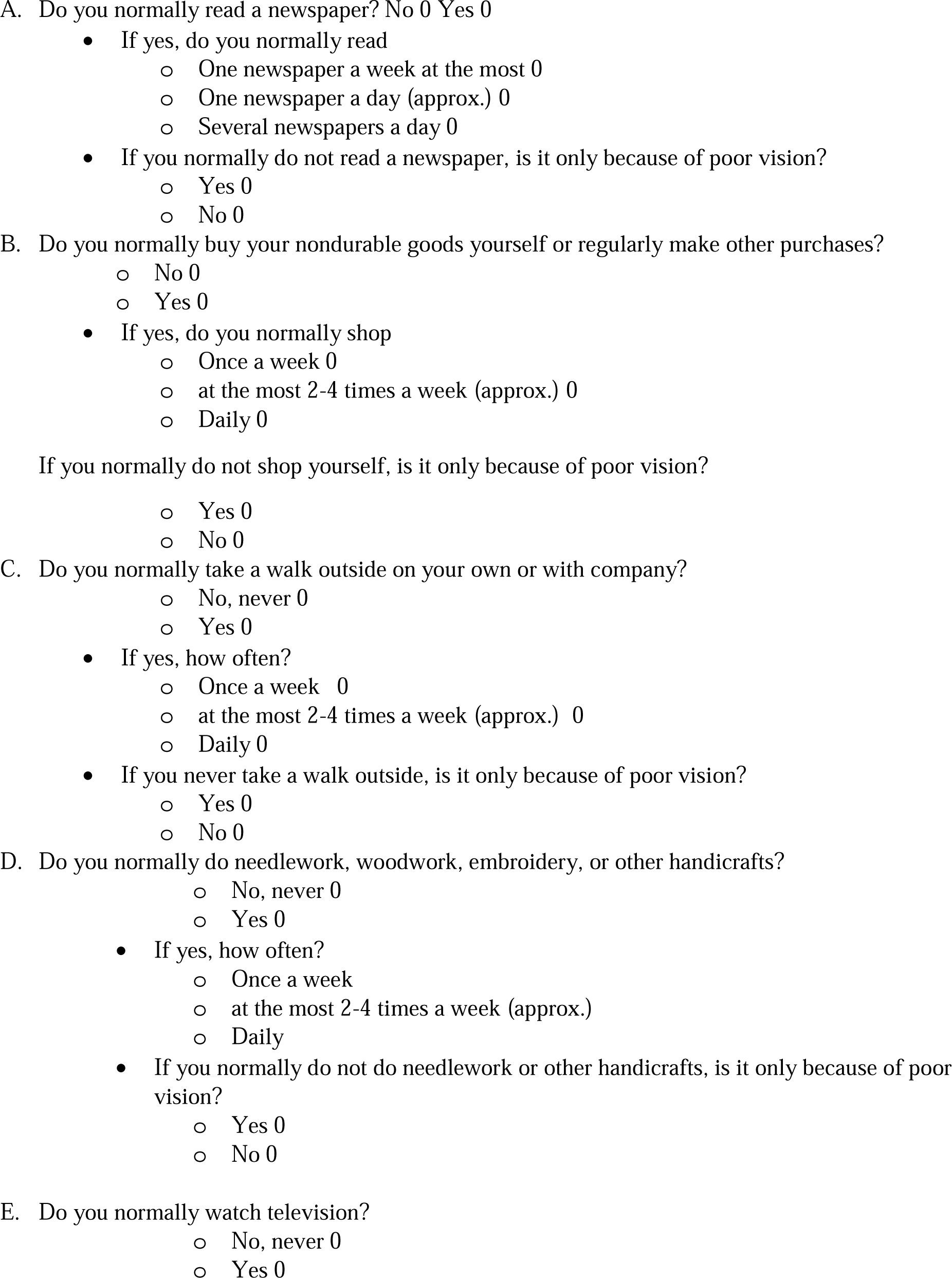
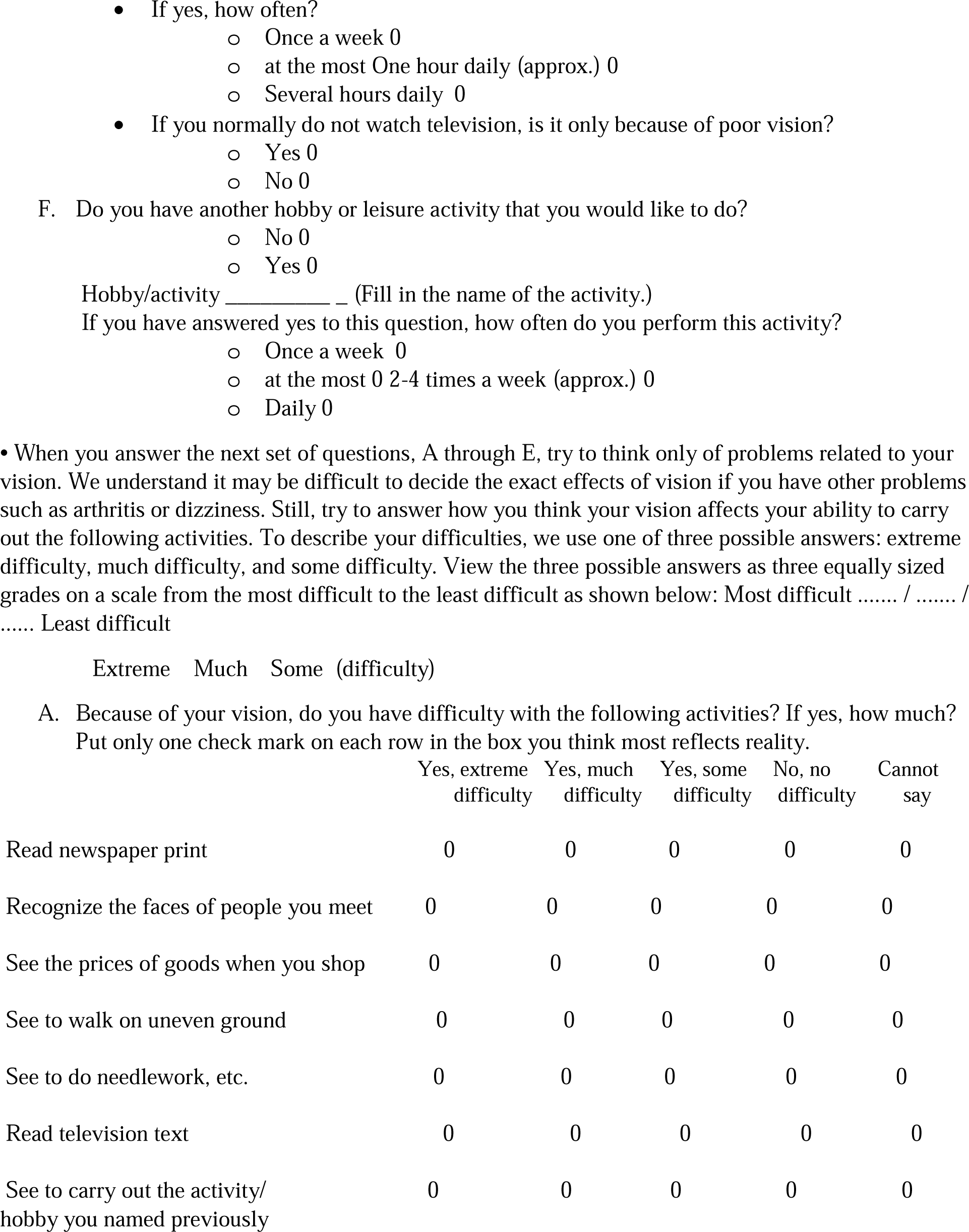
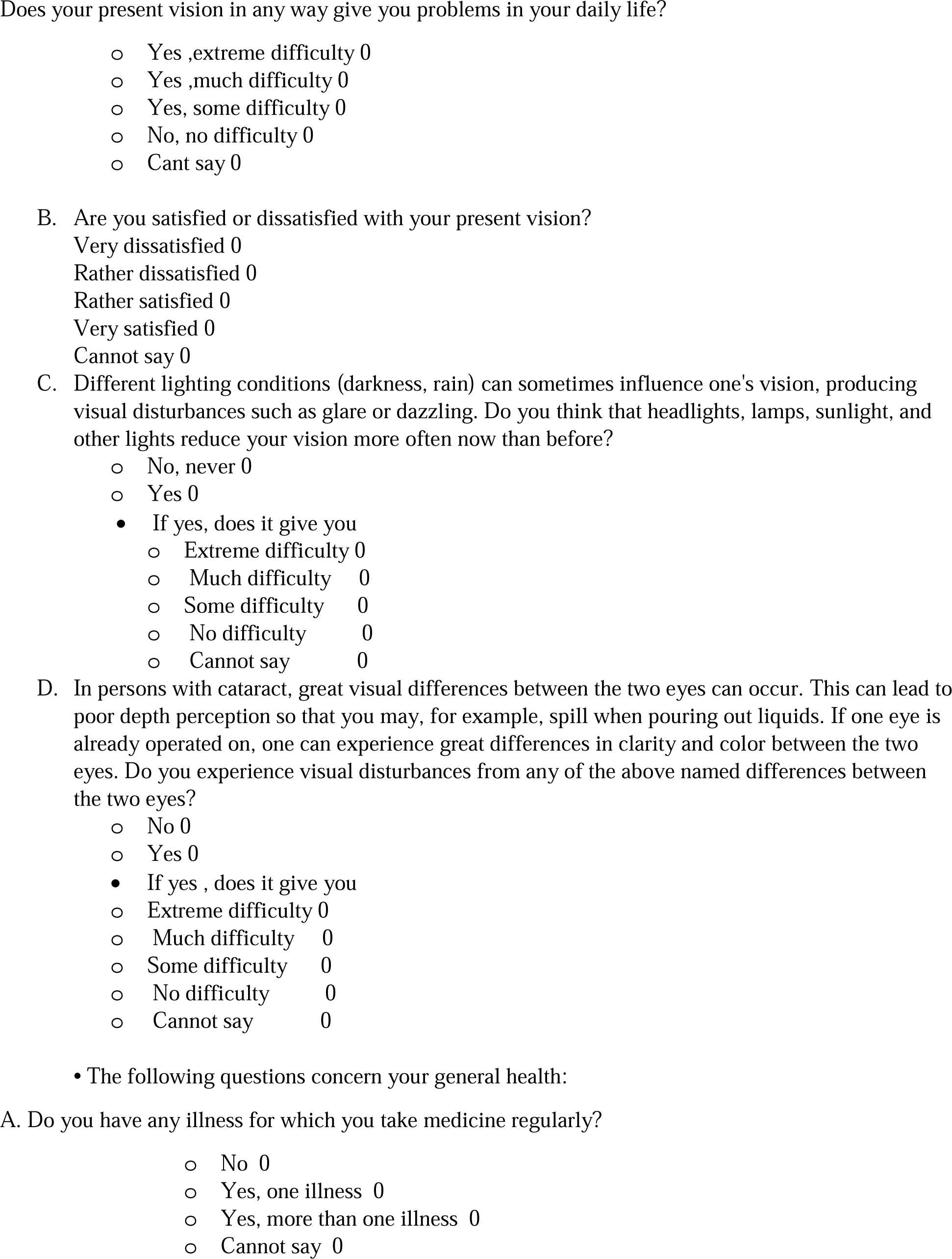
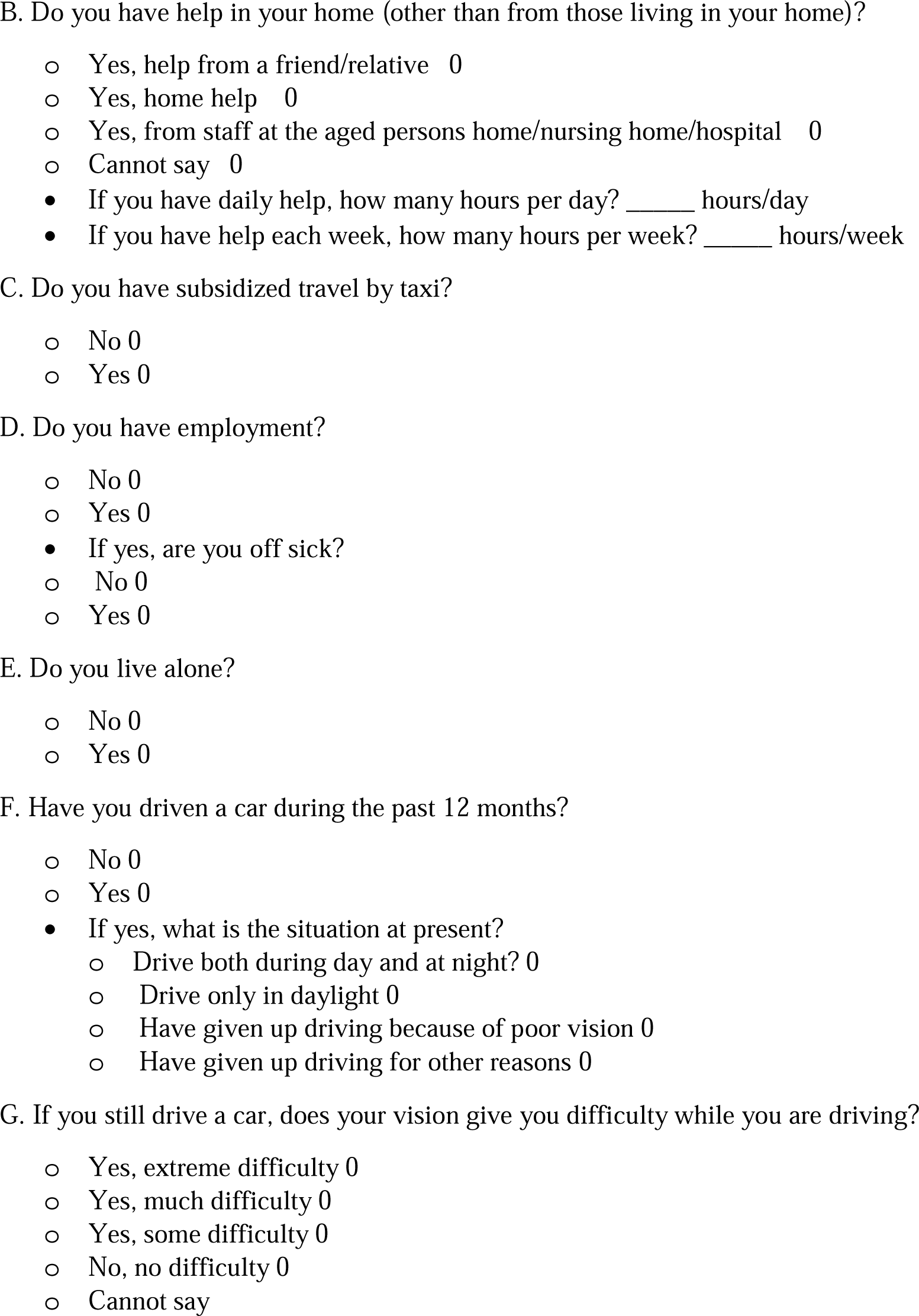

## Declaration

The manuscript has been read and approved by all authors and the authors declare no competing interests/conflict of interest

## Contribution Details

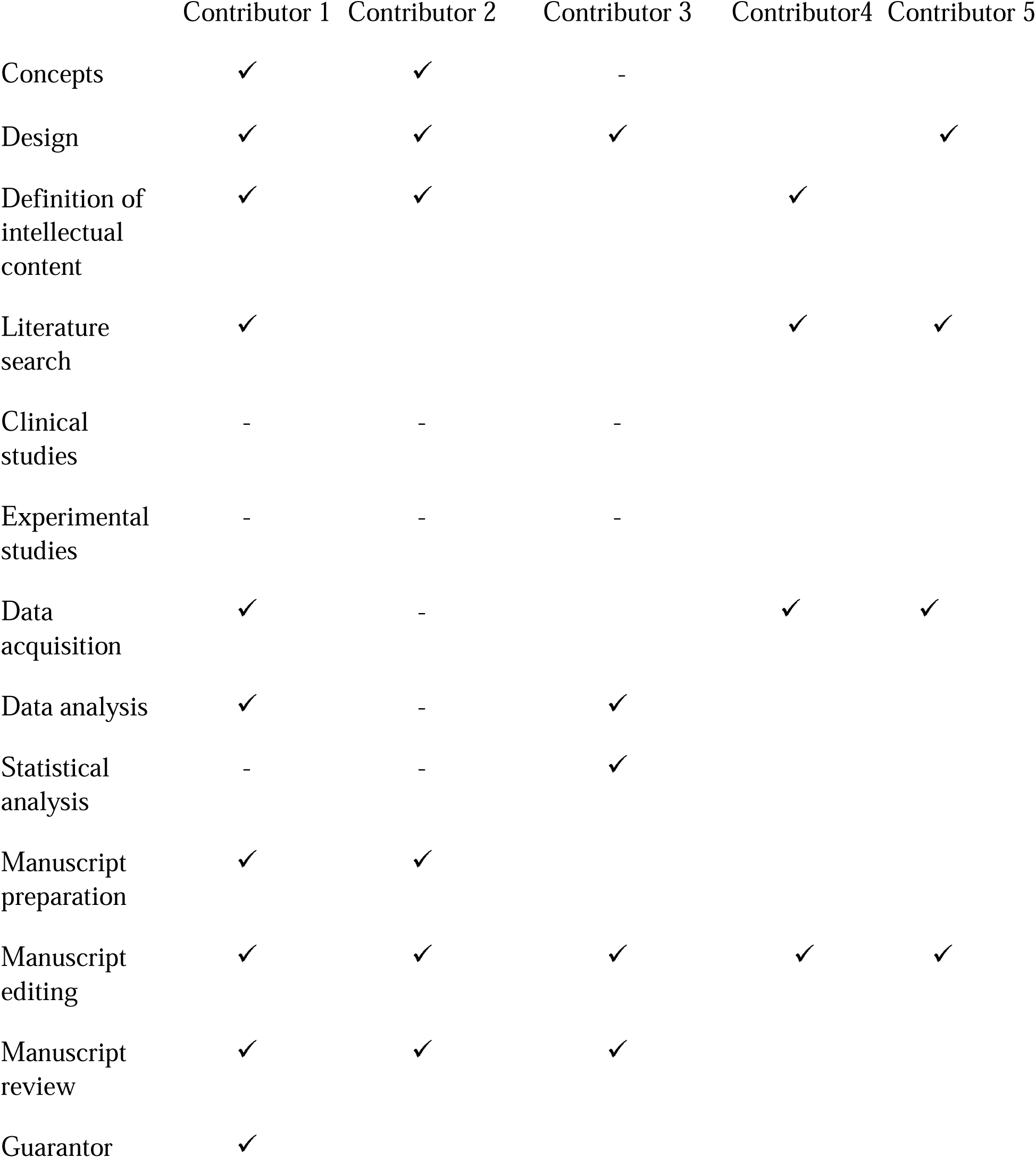

